# Home-based tDCS as an add-on to digital cognitive behavioral therapy application (dCBT app) in adults with ADHD: A sham-controlled randomized pilot study

**DOI:** 10.64898/2026.05.21.26353771

**Authors:** Katharina Kerkel, Andreas Reissmann, Laura Treml, Martin Schecklmann, Gitta Jacob, Mirja Osnabruegge, Berthold Langguth, Stefan Schoisswohl

## Abstract

**Introduction:** Over 30% of adults with Attention-Deficit/Hyperactivity Disorder (ADHD) show an insufficient response to standard pharmacological treatments, which underscores the need for evidence-based alternative interventions.

**Methods:** In this sham-controlled study, 30 adult outpatients with ADHD were randomized to 12 weeks of active or sham transcranial direct current stimulation (tDCS) as add-on to a digital cognitive behavioral therapy application (dCBT app). Participants received either active (2 mA, 20 min/day, 5 days/week) or sham tDCS with anodal (left) and cathodal (right) stimulation applied over the dorsolateral prefrontal cortex (DLPFC). In parallel, access to the dCBT app was provided for three months. ADHD symptoms were measured before and after treatment and after a three-month follow-up using the Adult Self-Report Scale (ASRS v1.1).

**Results:** All scales showed an improvement over time with medium-to-large within-subjects effects (Cohen’s d: -.48 to -.75), irrespective of group allocation. Two additional sensitivity analyses including (1) participants with ≥75 % of planned (sham)-tDCS sessions and (2) those who logged into the dCBT app on at least 5 days (median split) confirmed results. Response was observed in 1/15 (6.7%) of the tDCS group and 2/15 (13.3%) of the sham-tDCS group, with no difference between groups (*p* = .543, φ = -.111). Compliance to (sham-)tDCS was high. tDCS usability was rated marginally lower in the tDCS group.

**Conclusions:** tDCS as an add-on therapy could not produce additional improvement in ADHS symptoms. The results are discussed in terms of contextual and patient-related aspects.

**ClinicalTrials.gov Identifier:** **NCT06766214**.

## Introduction

### Attention-Deficit/Hyperactivity Disorder (ADHD)

Attention-Deficit/Hyperactivity Disorder (ADHD) is a neurodevelopmental disorder that commonly begins in childhood (Cherkasova et al., 2013) and frequently persists into adulthood, with up to 70% of affected individuals continuing to experience clinically relevant symptoms later in life (Cortese et al., 2025). With a prevalence of more than 5% among children and adolescents under the age of 18 (Faraone et al., 2021), ADHD represents one of the most common psychiatric conditions of childhood. Clinically, the disorder is characterized by persistent symptoms of inattention and/or hyperactivity that correspond to the diagnostic categories F90.0 or F98.8 in the ICD-10 classification (Doernberg & Hollander, 2016). The diagnostic process is often challenging due to pronounced heterogeneity in symptom presentation, developmental trajectories and associated impairments (da Silva et al., 2023). In adults, previous literature suggested a prevalence of approximately 2.5% (Simon et al., 2009), while more recent reviews report pooled prevalence rates of 3.1%, with the inattentive subtype being the most frequent, followed by the hyperactive and the combined type (Ayano et al., 2023). Although historically conceptualized as a predominantly male disorder, accumulating evidence shows that ADHD affects females to a comparable extent, albeit with differing symptom profiles and clinical trajectories (Faheem et al., 2022). Across lifespan, ADHD is associated with marked functional impairments across occupational, social and daily life domains (Bellato et al., 2025). Notably, over 60% of individuals with ADHD present at least one comorbid psychiatric disorder such as anxiety disorders, mood disorders or substance use disorder, underscoring the clinical complexity of the condition (Seo et al., 2022; da Silva et al., 2023). Despite the availability of effective medications, approximately 30% of adults with ADHD show an insufficient response to standard pharmacological treatments (Torgersen et al., 2008). This treatment gap underscores the need for accessible evidence-based alternative interventions that can complement medication in routine clinical care and address the multifaceted neurocognitive impairments associated with this disorder (Basiri & Hadianfard, 2023).

### Cognitive behavioral therapy (CBT)

Albeit inferior to pharmacological treatment (Philipsen et al. 2015), cognitive behavioral therapy (CBT) has demonstrated efficacy in reducing ADHD symptom severity and the associated functional burden in adults and is recommended as a standard treatment for ADHD patients (Tourjman et al., 2022). CBT has been shown to alleviate internalizing symptoms of ADHD, to support emotional regulation and helps patients to develop coping strategies in order to compensate for core ADHD symptoms (López-Pinar et al., 2020; Liu et al., 2023). In some studies, dialectical behavior therapy (DBT) has also been used as a potential adjunctive or alternative treatment for adults with ADHD (Basiri & Hadianfard, 2023). Originally developed to address chronic suicidal behavior and borderline personality disorder, DBT targets core difficulties such as impulsivity and deficits in emotional regulation (Basiri & Hadianfard, 2023). Given the overlap of these behavioral features between borderline personality disorder and ADHD, DBT has been adapted to meet the specific therapeutic needs of adults with ADHD, thus providing a framework for use in clinical settings (Philipsen et al., 2010). Nevertheless, access to clinicians with expertise in adult ADHD remains limited, leaving many patients without feasible treatment options (Schneider et al., 2023). Digital cognitive behavioral therapy applications (dCBT apps) have demonstrated robust effectiveness across a range of psychiatric conditions. A randomized controlled trial, in which patients were given three-month access to a dCBT app (*attexis*®), demonstrated significantly lower ADHD symptom severity in the intervention group compared with a waiting list control group, both immediately after a three-month treatment period and three-month follow-up (D’Amelio et al., 2026).

### Transcranial direct current stimulation (tDCS)

Another potential treatment approach involves non-invasive brain stimulation techniques such as transcranial direct current stimulation (tDCS). tDCS is capable of modulating cortical excitability (Salehinejad et al., 2020) via weak constant direct currents (typically 1–2 mA), which are delivered via scalp electrodes. These currents induce shifts in resting membrane potential that can either enhance or inhibit neuronal activity depending on electrode polarity. In the context of ADHD, stimulation is typically applied over the dorsolateral prefrontal cortex (DLPFC) (Bernadotte & Zinchenko, 2025; Courrèges et al., 2025), with anodal stimulation typically targeting the left DLPFC (F3, corresponding to the 10-20 EEG system) and cathodal stimulation targeting the right DLPFC (F4), aiming to increase and decrease cortical excitability, respectively (Nitsche & Paulus, 2000). In healthy adults, DLPFC stimulation has been shown to be effective across a range of attentional tasks, as demonstrated in a meta-analysis by Yadollahpour, Asl, and Rashidi (2017), indicating that tDCS can modulate attentional control. Building on these findings, in clinical studies, repeated DLPFC-targeted tDCS over several weeks has been shown to induce neuroplastic changes and to ameliorate core symptoms in adults with ADHD (Kuo, Chen & Nitsche, 2017; Bernadotte & Zinchenko, 2025; Courrèges et al., 2025). tDCS studies for ADHD have been performed with treatment applied both in the clinic and at home. **In-clinic studies** have demonstrated beneficial effects on impulsivity and attention. Allenby et al. (2018) reported reduced impulsivity in adults with ADHD following two blocks of three sham or active tDCS sessions targeting the left DLPFC, with stimulation order randomized (counterbalanced) and separated by a two-week interval, yielding medium effect sizes (d = 0.5), which were no longer evident at the 3-day post-stimulation follow-up. Cachoeira et al. (2017) observed significant symptom improvements in a sham-controlled design following five days of 2 mA tDCS (20 min/day), with the anode placed over the right DLPFC and the cathode over the left DLPFC, yielding large immediate post-stimulation effect sizes (d ≈ 0.6–0.9), which tended to gradually diminish over repeated follow-ups up to four weeks after stimulation. By contrast, Westwood et al. (2023) conducted a randomized, sham-controlled trial and found that three weeks of 1 mA tDCS over the right inferior frontal cortex combined with cognitive training did not yield significant effects. A **home-based** study further supports the efficacy of repeated DLPFC stimulation: Leffa et al. (2022) conducted a randomized, double-blind trial including 64 adults with ADHD not taking stimulant medication, showing that daily 30-minute home-based tDCS sessions over four weeks (2 mA; anodal right and cathodal left prefrontal stimulation) significantly improved symptoms of inattention (d = 1.23) without serious adverse events. Taken together, these findings indicate that repeated DLPFC-targeted tDCS, whether delivered in-clinic or at home, holds promise for alleviating attentional deficits and impulsivity in adults with ADHD.

### Combined treatment

To date, only a few studies have explored the combination of tDCS and cognitive training, as such a multimodal approach could yield synergistic effects and improve treatment outcomes (Spagnolo et al., 2020): An in-clinic study by Basiri and Hadianfard (2023) evaluated the effects of 1.5 mA tDCS administered alongside DBT in adults with ADHD. Participants received anodal and cathodal stimulation over F3 and F4, respectively, with each session lasting 20 minutes across 10 sessions and a 72-hour interval between sessions. The study found that tDCS alone did not reduce hyperactivity and DBT alone did not improve attentional deficits, whereas the combination of tDCS and DBT produced significant reductions across all ADHD symptoms. Importantly, the intervention was feasible and safe.

The results by Basiri & Hadianfard (2023) indicate that although combined interventions may offer benefits, the current evidence base regarding multimodal therapy approaches remains limited.

### Research Question

To the best of our knowledge, research on combined treatment approaches in adult ADHD remains scarce. While an in-clinic study by Basiri and Hadianfard (2023) demonstrated that combined interventions can improve ADHD symptomatology across multiple domains, the efficacy of such approaches has yet to be systematically investigated. Further, no study has yet investigated the efficacy of a combined treatment of a dCBT app and a neuromodulatory device in a home-based setting within a naturalistic sample. Hence, the objective of the present study was to further investigate home based tDCS as add-on to a dCBT app (*attexis*®, D’Amelio et al., 2026) within a sham-controlled randomized design to assess feasibility, compliance and changes in ADHD symptom severity following a 3-month treatment phase as well as a 3-month follow-up period. Insights gained aim to contribute to evidence base for accessible multimodal care in adults suffering from ADHD. Therefore, the following main research question was investigated: Can tDCS augment the therapeutic effects of an established dCBT app on symptom severity in adult ADHD?

## Methods and materials

### Subjects and study design

All methodological procedures for the implementation of the present study were approved by the ethics committee of the University of Regensburg (24-3862-101). The trial was registered at the U.S. National Institutes of Health Database (www.clinicaltrials.gov) accessible with the identifier code NCT06766214. The study presented here is a randomized, single-blinded and sham-controlled clinical trial conducted at the Center for Neuromodulation of a tertiary psychiatric hospital in Regensburg, Germany. Each participant was provided with two home-based therapies: a dCBT app and tDCS (verum or sham). Please find a detailed description of both therapies below.

A total of 33 German-speaking participants were recruited from September 2024 to August 2025 to achieve a final sample of *N* = 30. Three participants, all assigned to the sham group, discontinued the study: one withdrew consent spontaneously before study initiation and two withdrew after completing 8 and 16 sessions, respectively, citing lack of interest in continuing. No serious adverse effects occurred.

Recruitment took place via multiple strategies: (1) a poster announcing the study was published on the website of the Center for Neuromodulation (Regensburg, Germany), (2) psychotherapeutic ADHD-self-support-groups were contacted to inform potential participants and (3) psychiatric and psychotherapeutic colleagues who regularly treat ADHD patients were notified about the study. Additionally, four participants who had been treated for depression with antidepressant transcranial magnetic stimulation (TMS) and were considered remitted during their final TMS consultation were also included in the study as they suffered from a comorbid ADHS diagnosis. As soon as a potential participant expressed interest in the study, a telephone call with the study team took place to explain the course of the study and to assess eligibility according to the predefined inclusion and exclusion criteria (see **Table 1**).

**Table 1.**
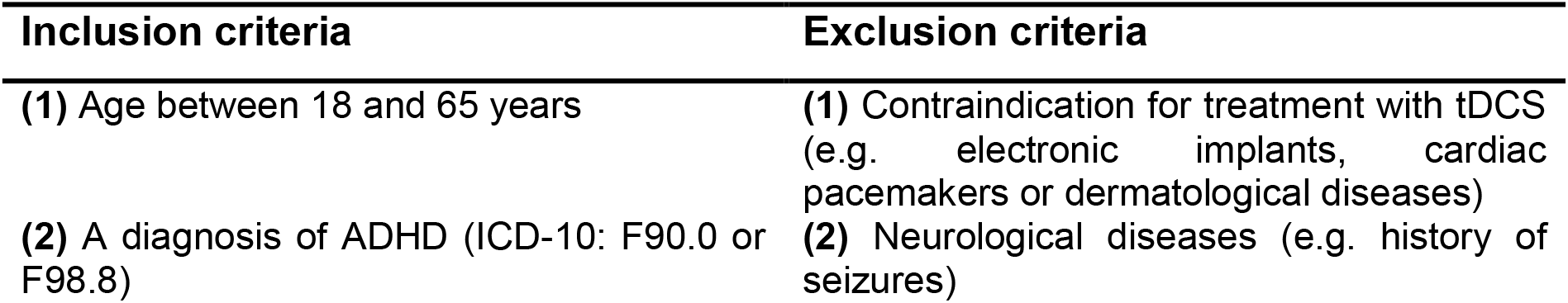
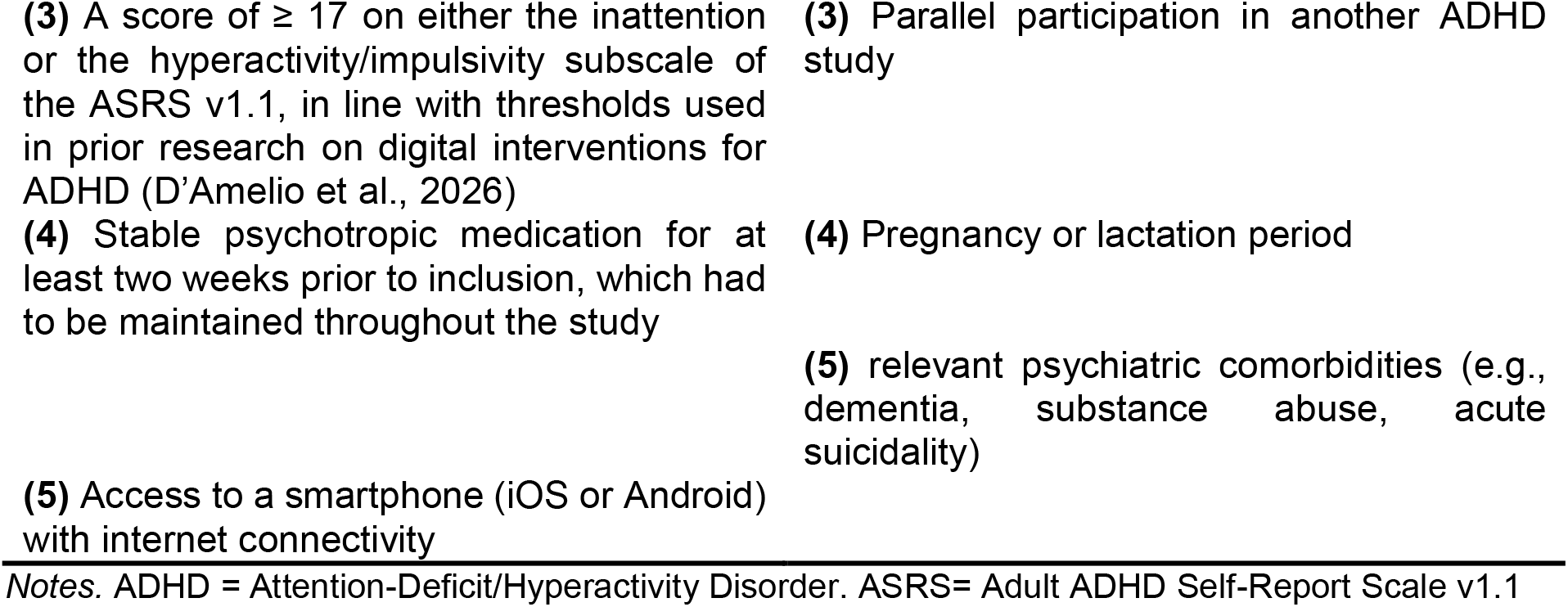
In- and Exclusion criteria for study participation.

Participants meeting all criteria were invited to the informed consent session on site. All participants were required to present documented verification of their ADHD diagnosis, such as an official medical certificate or a letter issued by their healthcare provider. Diagnosis was confirmed at the baseline visit by clinical assessment and the ASRS version 1.1. During this session, all participants received detailed information regarding study objectives, provided written informed consent and completed all baseline questionnaires (see Assessment). Afterwards, all participants were instructed on the correct use of the tDCS device itself. For the duration of the treatment, all participants were provided with a small medical kit containing the stimulator, consumables, 6 x 250 ml 0.9% sodium chloride solution, a 20 ml syringe and an adapter for the syringe to get easy access to the sodium chloride solution. As a last step, after the participants left the informed consent session, all participants were assigned to either the verum group (dCBT app + tDCS) or sham group (dCBT app + sham-tDCS). The form of stimulation (verum or sham) on the device itself was selected in the Sooma portal (see below for a detailed description of the stimulator). Computerized randomization was conducted prior to the start of the study (https://www.random.org/lists/). Therefore, initially, pseudonymized codes containing ascending numerical identifiers were generated. As participants enrolled over time, each participant was assigned the next available code according to their order of appearance. Group allocation was then determined based on the pre-randomized sequence of these pseudonymized codes, ensuring random assignment independent of the timing of enrollment. All participants got access credentials for all study platforms via e-mail on the Friday of the inclusion week. Treatment phase commenced the following Monday. (Sham-)tDCS sessions were administered daily (2mA, 20 minutes per session). During the first week of treatment, daily video consultations via the CLICKDOC software (version 5.9.1, La-Well Systems GmbH; a clinically approved platform) ensured correct device operation and provided guidance, if needed. Once participants demonstrated independent and correct use of the tDCS device, treatment continued without further accompaniment. Nevertheless, every participant was allowed to contact the investigators at any time in case of side effects or other questions. For this purpose, important contact details were provided to all participants. Patients were able to access the dCBT app at their convenience throughout the 12-week study period. On days when participants completed both interventions, the order of administration (tDCS followed by login to the dCBT application, or vice versa) was not prescribed and could be chosen freely. After three months of treatment, a final meeting was arranged with the participants by email or telephone, during which they returned the kit, filled out questionnaires, were asked for open feedback and were informed about their group assignment. Three months later, all participants were contacted via email and asked to complete a follow-up questionnaire. **Figure 1** illustrates the course of the study for one participant.

**Figure 1.**
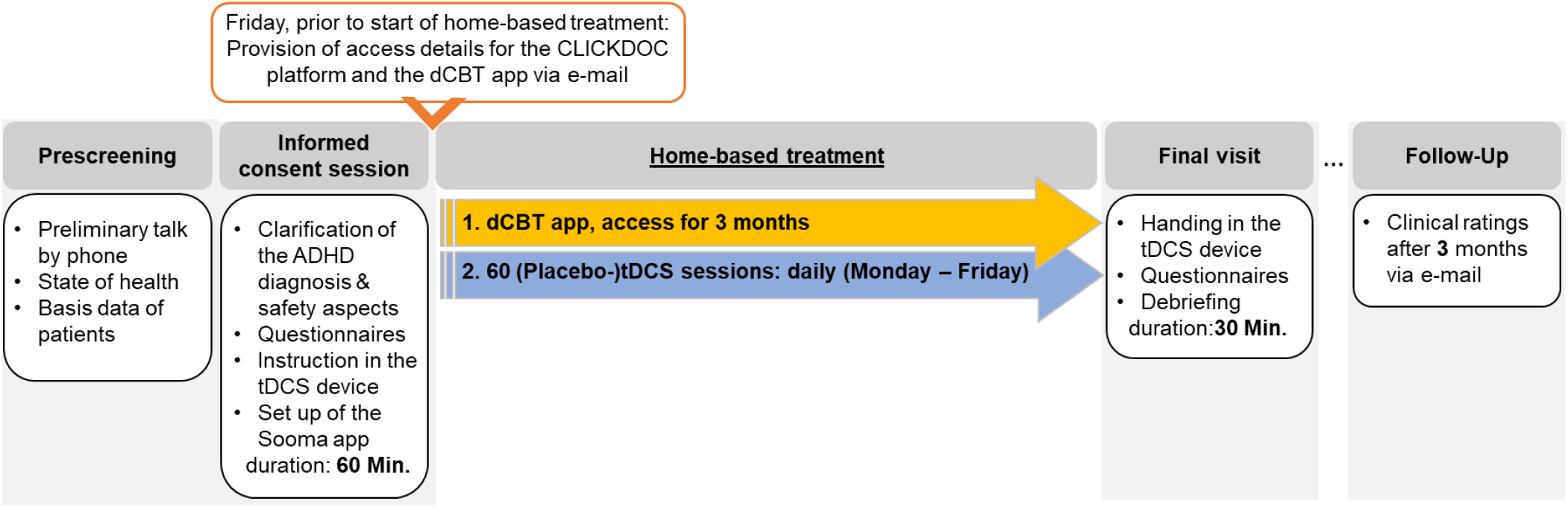
Course of the study. The figure illustrates the treatment schedule for one participant using the dCBT app combined with (sham-)tDCS. After eligibility was confirmed in a preliminary phone screening, the first on-site session took place. Participants received study information, the (sham-)tDCS device and login data for CLICKDOC and the dCBT app (sent on the respective Friday before treatment started on Monday). During the intervention, patients self-administered daily prefrontal (sham-)tDCS at 2 mA for 20 minutes and had access to the dCBT app for three months. The final on-site session included returning the device, study debriefing and information about group assignment. BKH = Bezirksklinikum Regensburg: tertiary care hospital in Regensburg, Germany. (Sham-)tDCS = (Sham-) transcranial direct current stimulation. ADHD = Attention-Deficit/Hyperactivity Disorder. dCBT app = digital cognitive behavioral therapy application (*attexis***®**)

### (Sham-) transcranial direct current stimulation (tDCS)

All stimulations were performed with Sooma Duo tDCS devices^TM^ (Sooma Oy, Helsinki, Finland) (see **Figure 2**). The devices include built-in safety mechanisms such as continuous impedance monitoring and automatic termination of stimulation in case of insufficient electrode contact or high impedance. The delivered current is controlled to match the predefined stimulation parameters and output as well as ramp rates are restricted to established safety limits. Regular impedance checks are performed throughout the session. Based on the study by Basiri & Hadianfard (2023), all participants were treated over the DLPFC. In the tDCS group, in each session a current of 2 mA was applied for 20 minutes with the anode placed over the left DLPFC, corresponding to F3 in the international 10–20 EEG system, and the cathode placed over the right DLPFC (F4). Sham-tDCS was implemented using a standard ramp-up/ramp-down procedure to mimic the sensation of active stimulation. The current was ramped up at 0.1 mA/s to the target intensity of 2 mA, followed by an immediate ramp-down at −0.1 mA/s back to 0 mA. No current was delivered thereafter, except for brief impedance checks performed every 5 minutes, during which a current of 0.1 mA was applied for 2 seconds. A cap with pre-punched holes at F3 and F4 facilitated correct placement of the electrodes (Large/Medium/Small ComfoCap, Sooma Medical, Sooma Oy, Helsinki, Finland). For each participant, the researcher initially determined the appropriate cap size by eyesight. A pair of round rubber electrodes with dimensions of 25cm^2^ (ComfoTrode, Sooma Medical, Sooma Oy, Helsinki, Finland) were used to apply the current. Before the treatment cap was placed on the subject’s head to initiate the treatment, tDCS electrode sponges (ComfoPad, Sooma Medical, Sooma Oy, Helsinki, Finland) were soaked in a 0.9 % sodium chloride solution to lower the physiological skin resistance, which were then placed into the electrodes.

**Figure 2.**
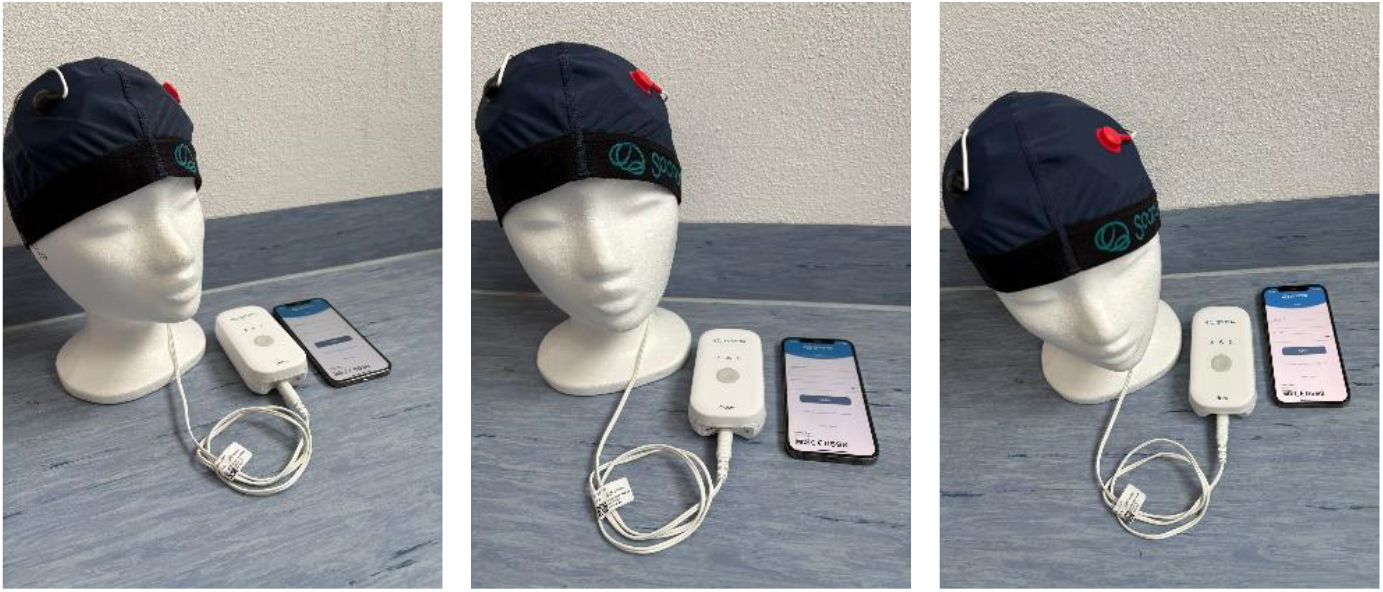
Sooma Duo tDCS devices™ (Sooma Oy, Helsinki, Finland) (own source). These images illustrate the (sham-)tDCS setup used in the study, including the electrode cap, stimulator and the Sooma app installed on a smartphone. The second intervention in the study was a digital cognitive behavioral therapy application (dCBT app, *attexis*®; here not shown).

All participants were able to start the treatments using a dedicated Sooma application, which can be installed on all iOS or Android devices. The study coordinators got access to the Sooma medical Portal (https://portal.soomamedical.com/login) in order to be able to register each study participant. The portal enabled the team to register each participant, program each device and monitor compliance. If a participant missed treatments on three consecutive days, the respective participant was reminded via e-mail to continue the treatment.

### Attexis® (digital cognitive behavioral therapy application, dCBT app)

The dCBT app was created and is owned by GAIA AG (Hamburg, Germany) and was developed by a multidisciplinary team of psychologists, physicians, software engineers and communication specialists as a fully self-guided digital health intervention for adults aged 18 years and older. The dCBT app is based on cognitive behavioral therapy (CBT) and mindfulness principles (D’Amelio et al., 2026) and is designed for independent use without professional guidance, allowing users to access the program online at their convenience within a predefined time window.

It provides a structured, home-based approach to delivering evidence-based psychosocial care while addressing barriers such as limited access to specialized clinicians. The application delivers personalized psychoeducational content and therapeutic exercises tailored to individual needs and preferences related to ADHD symptoms. A central feature is the use of “simulated dialogues,” in which users interact with brief text passages by selecting response options that guide the flow of information and individualize the experience. To support compliance and motivation, the application incorporates adaptive content presentation and automated reminders: participants who did not log into the application for more than two weeks received an email reminder.

### Assessment

The Adult ADHD Self-Report Scale (ASRS, version 1.1), which is a self-administered questionnaire designed to assess ADHD symptoms in adults, was filled out by participants before and after treatment and as part of the follow-up survey. It includes 18 items covering the two main symptom domains of inattention and hyperactivity/impulsivity, with participants rating how often they experience each symptom on a five-point scale from “never” to “very often.” The ASRS v1.1 total score is computed by summing up all 18 items, resulting in a range between 0 and 72. Subscale scores can be calculated for both subscales. Higher scores indicate greater symptom severity. The ASRS v1.1 has been validated in various adult populations and demonstrates good reliability and internal consistency (Mörstedt, Corbisiero & Stieglitz, 2016), making it a widely used tool in both research and clinical practice for assessing symptom severity (Carlucci et al., 2017).

To investigate the subjective patient experience with the (sham-)tDCS devices, all participants filled out the User Experience Questionnaire at the final visit (UEQ; Schrepp, Thomaschewski & Hinderks, 2017). The questionnaire is divided into six scales, which measure usability as well as user experience aspects (7-step Likert-Scale ranging from -3 to +3). The six scales include Attractiveness (Overall impression of the product), Perspicuity (Is it easy to get familiar with the product?), Efficiency (Can users solve their tasks without unnecessary effort?), Dependability (Does the user feel control of the interaction?), Stimulation (Is it exciting and motivating to use the product?) and Novelty (Is the design of the product creative?) (https://www.ueq-online.org/; access: 2025-03-21). Further, beginning with the ninth enrolled participant, participants’ guesses regarding their group assignment (tDCS vs. sham-tDCS) were systematically collected at the final visit. In addition, open-ended feedback on both the dCBT app and the tDCS intervention was obtained to assess user experience and satisfaction.

### Statistical analysis

All statistical analyses were conducted using SPSS version 31.0.0 (IBM SPSS, Chicago, IL). Group differences (tDCS vs. sham-tDCS) were calculated by chi-square tests of independence or Student t-tests. For the analysis of the course of the ADHD symptoms, three mixed analyses of variance (ANOVA) were calculated: for the total sum score as well as the subscales hyperactivity and inattention with time as within factor (2 levels: before and after treatment) and group as between factor (2 levels: tDCS and sham-tDCS). In addition to partial η^2^ for ANOVA effects (see below), Cohen’s d (Cohen, 1988) was calculated to quantify within-subject pre–post changes using pooled standard deviations. In case of a significant interaction, a post-hoc t-test was performed. Further, to assess the robustness of the findings, two sensitivity analyses were conducted. In the first sensitivity analysis, only participants who completed ≥75 % of the planned (sham-)tDCS sessions were included, in the second analysis only those participants who logged into the dCBT app on at least five days (corresponding to the median number of days on which participants used the app) were included into the analyses. Accordingly, the mixed ANOVAs were repeated in the high-compliance subgroups. As in D’Amelio et al. (2026), responders were defined as participants who showed a ≥30% reduction in the ASRS total sum score from before to after treatment (Buitelaar, Montgomery, & van Zwieten-Boot, 2003). Since email-based follow-up resulted in >50% missing data due to non-response, the follow-up results are reported for descriptive purposes only. Compliance with the (sham-)tDCS treatment was assessed as the percentage of completed sessions relative to the number of programmed sessions. Compliance with the dCBT app was defined as the number a participant logged into the application. The number of days a participant logged into the application was monitored by the developer of the dCBT app. Differences in the distribution of guessed group assignments between study groups were analyzed using Fisher’s exact test. The mean values of the UEQ were analyzed with a preset Excel sheet, which is open to the public (ueq-online.org), by Schrepp, Thomaschewski and Hinderks (2017). Level of significance was set to *p* <.05. As described above, for effect sizes we used Cohen’s *d* (Cohen, 1988), partial η^2^ or the phi-coefficient (03C6). By convention (Cohen, 1988), effect sizes are divided in small (*d* = 0.2; partial η^2^ = 0.01; φ = 0.1), medium (*d* = 0.5; partial η^2^ = 0.06; φ = 0.3), and large effects (*d* = 0.8; partial η^2^ = 0.14; φ = 0.5).

## Results

### Demographic and clinical characteristics

All demographic and clinical characteristics of the enclosed participants are provided in **Table 2**. The mean age was approximately 31.47 (± 9.70) years, with 53% reporting female sex. Most participants were living either alone (47%) or with a partner (40%), had A-Levels (73%) and were working full-time (47%). 63% were taking psychotropic medication, mostly psychostimulants.

**Table 2.** Demographic and clinical data of the present sample.

|  | tDCS<br>(*n* = 15) | sham-tDCS<br>(*n* = 15) | Statistics for group<br>comparisons |
| --- | --- | --- | --- |
| **General variables** |  |  |  |
| **sex: m/f** | 9/6 | 5/10 | $\chi^2(1) = 2.143, p = .143;$<br>$\phi = -.267$ |
| **age: M (SD)** | 30.47<br>(8.69) | 32.47<br>(10.83) | $t(28) = .558, p = .581, d =$<br>.20 |
| **age: range** | 20 – 49 | 20 – 59 |  |
| **Type of F9-Diagnosis (ICD-10)** |  |  |  |
| **F90.0/F98.8** | 14/1 | 11/4 | $\chi^2(1) = 2.160, p = .142;$<br>$\phi = -.268$ |
| **ADHD Medication<sup>1</sup> (e.g.**<br>**Medikinet, Elvanse): yes/no** | 8/7 | 11/4 | $\chi^2(1) = 2.222, p = .136;$<br>$\phi = -.272$ |
| **Questionnaire scores at**<br>**baseline: M (SD)** |  |  |  |
| **ASRS: total** | 50.40<br>(12.52) | 48.87<br>(8.34) | $t(28) = -.395, p = .696, d$<br>$= -.14$ |
| **ASRS: subscale attention** | 27.13<br>(6.37) | 26.73<br>(4.68) | $t(28) = -.196, p = .846, d$<br>$= -.07$ |
| **ASRS: subscale hyperactivity** | 23.47<br>(6.63) | 22.13<br>(4.36) | $t(28) = -.651, p = .520, d$<br>$= -.24$ |

Notes. <sup>1</sup> The time of intake (morning, noon, evening) was not systematically recorded. ADHD = Attention-Deficit/Hyperactivity Disorder. ASRS = Adult ADHD Self-Report Scale v1.1

Comorbid psychiatric disorders were present as follows: 30% of the sample had a depression diagnosis (ICD-10: F33.x), two participants had a generalized anxiety disorder (ICD-10: F41.1), two participants had a posttraumatic stress disorder (ICD-10: F43.1), one participant had a panic disorder (ICD-10: F41.0) and three participants had a comorbid personality disorder (ICD-10: F6).

## Efficacy

**Figure 3** provides mean changes in ADHD symptoms from before to after treatment. A mixed ANOVA regarding the total sum score of the ASRS revealed a significant effect of time (F_(1,28)_ = 21.236, *p* <.001, partial η^2^ = .431), corresponding to a medium within-subjects effect (Cohen’s d = - .66), No significant group effect (F_(1,28)_ = 0.038, *p* = .847, partial η^2^ = .001) or interaction (F_(1,28)_ = .555, *p* =.463, partial η^2^ = .019) was observed. Further, a mixed ANOVA regarding the inattention subscale also revealed a significant effect of time (F_(1,28)_ = 29.594, *p* <.001, partial η^2^ = .514), corresponding to a medium-to-large within-subjects effect (Cohen’s d = - .75). Neither a significant group effect (F_(1,28)_ = .001, *p* =.972, partial η^2^ < .001) nor a significant interaction between time and group was found (F_(1,28)_ = .493, *p* = .489, partial η^2^ = .017). A mixed ANOVA regarding the hyperactivity subscale also revealed a significant effect of time (F_(1,28)_ = 9.358, *p* =.005, partial η^2^ = .250), also corresponding to a medium within-subjects effect (Cohen’s d = -.48). No significant group effect (F_(1,28)_ = .152, *p* = .699, partial η^2^ = .005) nor a significant interaction between time and group (F_(1,28)_ = .570, *p* =.457, partial η^2^ = .020) were found.

**Figure 3.**
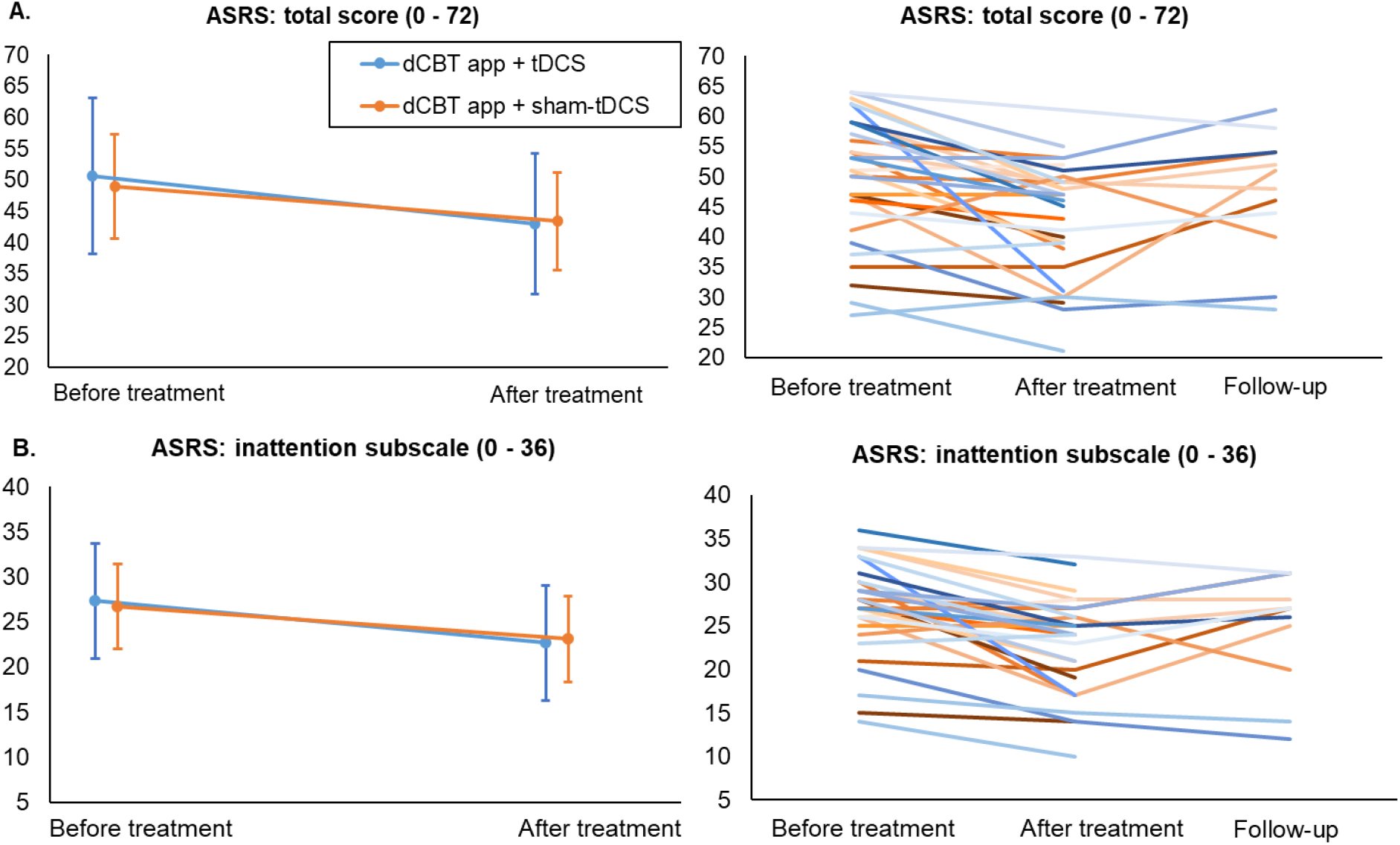
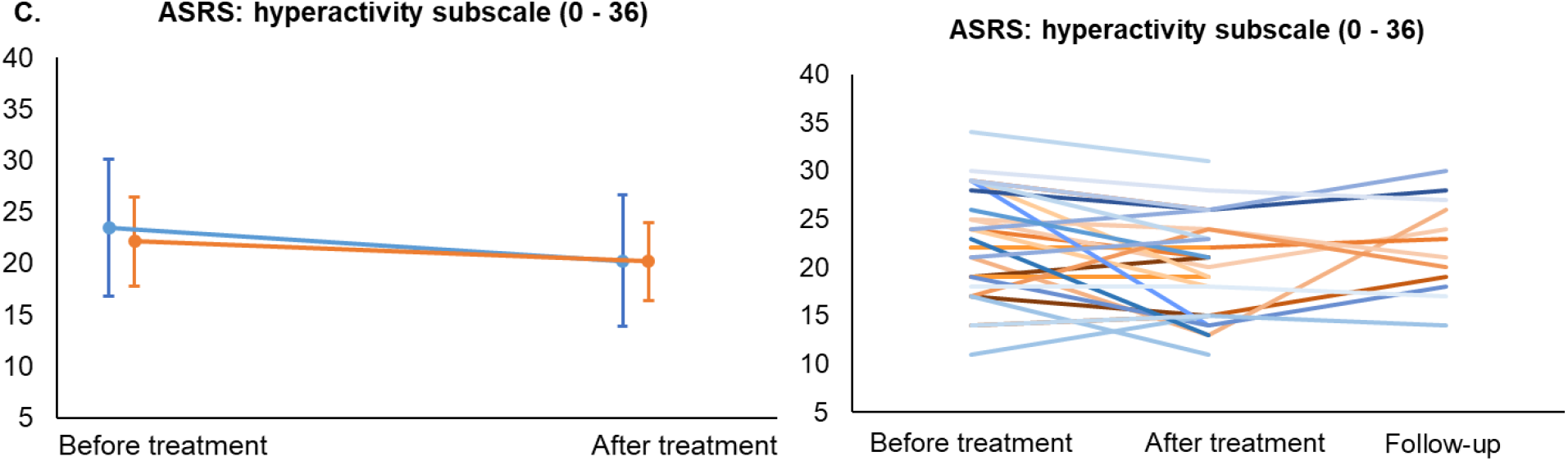
Progression of ADHD symptoms. Left side: Course of mean course of the ADHD scores (due to the fact that > 50% of the follow-up assessments were not returned, mean scores are shown only for the before and after treatment). Right side: Course of scores for each participant (three measurement time points). Orange: dCBT app + sham-tDCS; blue: dCBT app+ tDCS: (**A**.) the total score, (**B**.) the inattention subscale and (**C**.) the hyperactivity subscale of the ASRS. Error bars indicate SDs for the mean scores. ASRS = Adult ADHD Self-Report Scale v1.1

**Figure 4.**
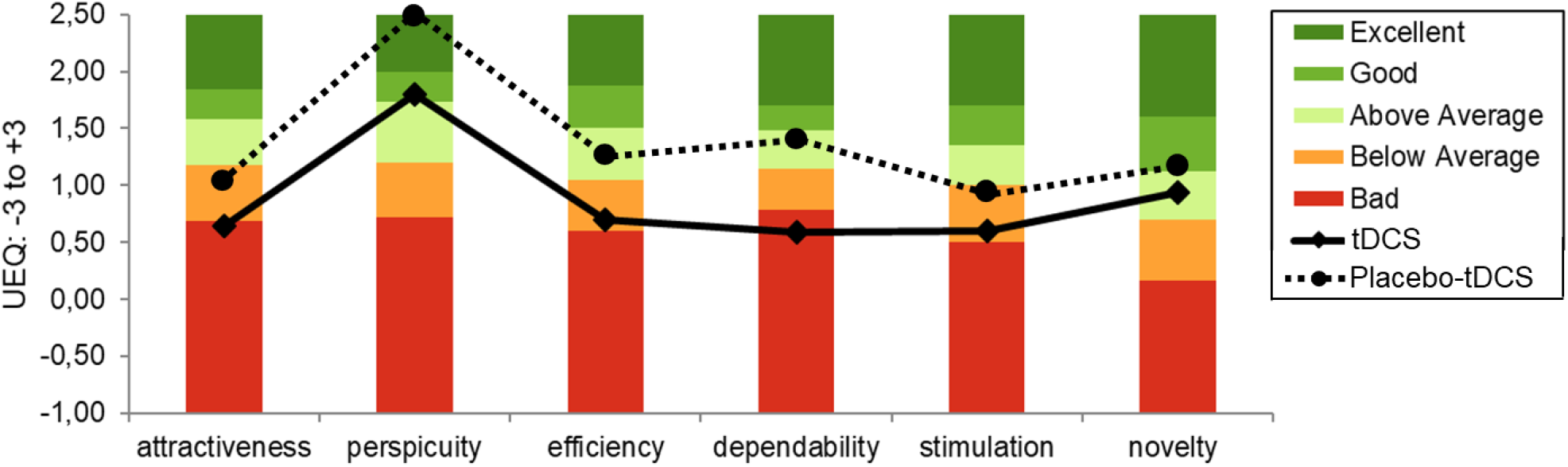
Mean evaluation of the user experience. This graph shows the mean scores and SDs of the six factors of the user experience questionnaire (UEQ) across the tDCS-group (♦) and sham-tDCS group (•) based on the open-source evaluation method by Schrepp, Thomaschewski and Hinderks (2017). Higher scores correspond to better evaluation.

For participants who completed ≥75 % of the planned (sham-)tDCS sessions (*n* = 17/30), mixed ANOVAs revealed significant main effects of time for the total ASRS score (F(1,15) = 9.778, *p* = .007, partial η^2^ = .395), the inattention subscale (F(1,15) = 12.108, *p* = .003, partial η^2^ = .447), and the hyperactivity subscale (F(1,15) = 6.787, *p* = .020, partial η^2^ = .312), all corresponding to medium within-subjects effects (Cohen’s d: -.52 to -.67). No significant between-subjects effects or interactions were observed for any scale (all *p*s > .323). For participants who logged into the dCBT app on ≥5 days (*n* = 17/30), mixed ANOVAs showed no significant main or interaction effects for the total score or subscales (all *p*s > .410).

After treatment, according to the ASRS total sum score, 6.7% of participants in the tDCS group (1/15) met the predefined responder criterion compared to 13.3% in the sham-tDCS group (2/15). The difference between groups did not reach statistical significance (χ^2^(1) = .370, *p* = .543; φ = -.111). Follow-up data were available for 12/30 participants (40%), 6 participants for each group.

### Compliance and User Experience

### 1. (Sham-)tDCS

#### 1.1. Compliance

Unfortunately, for one participant, fewer tDCS sessions than planned were programmed via the portal (56 sessions), while for two other participants, more sessions than planned were programmed (63 and 69 sessions). Another participant completed only 46 sessions, as the intervention was terminated earlier by the research team due to mild side effects (mild itching and skin redness), although the participant was willing to continue. However, since this participant completed more than 75% of the planned sessions, the participant was included in the analyses. An independent samples t-test revealed no significant difference between the tDCS (M = 80.83, SD = 14.75) and sham-tDCS group (M = 72.83, SD = 13.83) in the percentage of completed tDCS sessions (t(28) = -1.523, *p* = .137, d = -.56).

### 1.2. User Experience: Open feedback and UEQ

In an open-ended feedback, several participants reported that the daily requirement to complete the 20-minute intervention session helped to provide structure to their daily routine. Overall, patients in both groups reported that they managed the handling and operation of the device well. Throughout the course of the treatment, no patient raised any active inquiries regarding technical issues or difficulties in execution.

In the sham-tDCS group, 5/15 patients correctly identified their group assignment, whereas 8/15 guessed incorrectly; no data were obtained from two patients. In the tDCS group, 3/15 patients correctly identified their assignment and 5/15 guessed incorrectly; two patients reported no clear assumption and no data were obtained from five patients. A Fisher’s exact test revealed no significant association between group assignment and correct identification (*p* = .332).

Based on the evaluation method by Schrepp, Thomaschewski and Hinderks (2017), in the tDCS-group, the participants evaluated the tDCS-treatment as follows: the factor *perspicuity* (M = 1.80, SD = 0.68) was rated “good”. The factor *novelty* (M = 0.93, SD = 0.75) was rated “above average”. The factors *efficiency* (M = 0.70, SD = 0.98) and *stimulation* (M = 0.60, SD = 0.98) were rated “below average”. The factors *attractiveness* (M = 0.64, SD = 0.90) and *dependability* (M = 0.58, SD = 0.98) were rated “bad”.

The participants in the sham-tDCS group evaluated the treatment as follows: the factor *perspicuity* (M = 2.52, SD = 0.45) was rated “excellent”. The factors *efficiency* (M = 1.25, SD = 0.86) and *dependability* (M = 1.40, SD = 0.93) were rated “above average”. The factor *novelty* (M = 1.18, SD = 1.12) was rated “good”. The factors *attractiveness* (M = 1.04, SD = 0.99) and *stimulation* (M = 0.92, SD = 0.92) were rated “below average”.

Independent samples t-tests revealed that the factor perspicuity (t(28) = 3.422, *p* = .002, d = 1.25) as well as the factor dependability (t(28) = 2.341, *p* = .027, d = .86) were rated significantly better in the sham-tDCS group than in the tDCS-group. All other factors were rated the same (all *p*s >.114).

### 2. dCBT app

#### 2.1. Compliance

An independent samples t-test revealed no significant difference between the tDCS (M = 4.67, SD = 4.20) and sham-tDCS group (M = 6.20, SD = 3.86) in the number of days participants logged in the application (t(28) = 1.041, *p* = .307, d = .38). While login frequency was recorded, the extent to which participants engaged with the specific therapeutic content of the app could not be assessed.

#### 2.2. User Experience: Open feedback

With respect to the dCBT app, several participants reported that the interface included many colorful or playful elements, which in some cases made them feel that the application was less suited to their needs. Further, several patients reported they lacked a feature that would allow them to track their progress, like the number of sessions remaining. Some also indicated that the content did not provide new information or was partly too repetitive and in a few cases, this contributed to discontinuation of use. Nevertheless, a certain number of participants found the application helpful and were able to engage with it consistently, but with some completing the entire program within just two weeks.

## Discussion

Given prior evidence supporting the therapeutic benefits of the digital cognitive behavioral therapy application (dCBT app, *attexis*®) (D’Amelio et al., 2026), the present study investigated whether these beneficial effects could be enhanced through adjunctive home-based transcranial direct current stimulation (tDCS). Therefore, a single-blinded, randomized sham-controlled study was conducted, in which all participants received access to the application alongside either tDCS or sham-tDCS over the course of 12 weeks. Although a significant improvement in ADHD symptoms was observed, with effect sizes being broadly comparable to previous findings (ASRS total sum score: D’Amelio et al. (2026): Cohen’s d = −.85; present study: Cohen’s d = −.66), this effect was not moderated by group assignment, indicating that tDCS as an add-on intervention did not confer additional symptom improvement beyond the observed time effects. Sensitivity analyses including only participants who completed at least 75% of the (sham-)tDCS sessions confirmed these findings. Further sensitivity analyses applying a median split to define high-compliance dCBT app users showed that the significant main effect of time observed in the primary analysis including all participants was no longer present, likely due to the reduced sample size, which may be attributable to the dichotomization inherent in the median split.

With respect to **efficacy**, our results are not in accordance with a study done by Hadianfard and Basiri (2023), who also treated ADHD patients (*N* = 80) with tDCS as an add-on therapy for dialectic behavioral therapy (DBT) and could report beneficial effects from a combined treatment. The authors reported superior outcomes for a combined DBT and tDCS intervention compared to single-treatment conditions, with moderate-to-large between-group effects across symptom-related measures, while in the present study, we did not observe a significant group × time interaction, despite medium within-subjects effects (Cohen’s d ≈ -.60). A key methodological distinction lies in the control of medication status. Basiri and Hadianfard (2023) included exclusively non-medicated patients, thereby ensuring that treatment effects could be examined without pharmacological confounding. In contrast, in the present study, more than half of the participants in both groups were receiving ADHD medication. Given the well-established efficacy of pharmacotherapy in reducing ADHD symptoms (De Crescenzo et al., 2017; Ogundele & Ayyash, 2023; Radonjić et al., 2023; Veronesi et al., 2024), baseline symptom severity in our sample may already have been partially reduced. Consequently, the potential for detecting additional benefits attributable to a third therapy may have been limited, which could explain the absence of add-on effects in our study.

Another important distinction between our study and that of Basiri and Hadianfard (2023) lies in the treatment setting and the structure of intervention delivery. While Basiri and Hadianfard (2023) conducted both DBT and tDCS in the clinic and administered them always simultaneously, our interventions were delivered entirely in a home-based format with possible time windows to administer both therapies. Even though compliance with the (sham-)tDCS sessions was generally high, with participants completing 73% of sham-tDCS sessions and 81% of tDCS sessions, our protocol was nonetheless less structured than that of Basiri and Hadianfard. For instance, participants had a 24-hour window to complete a single (sham-)tDCS session before the device locked for the next session, resulting in variable treatment times. Further, access to the dCBT app was on the participants’ convenience and thus, participants were not required to follow a fixed schedule or set regular therapy times to complete all therapy interventions. Consequently, also simply due to the need to operate two separate therapy apps on the same smartphone (the dCBT app as well as the Sooma app to start the (sham-)tDCS session), simultaneous administration of the interventions was not possible (on days when participants completed both, the order of administration was not standardized). Concurrent application of both therapies might have led to different results, as a study by Besson et al. (2019) showed that applying tDCS during task performance results in greater sensorimotor cortex activation compared to sequential administration. In this context, Basiri and Hadianfard (2023) demonstrated that the simultaneous administration of DBT and tDCS was associated with improvements across overall ADHD symptomatology: While tDCS can enhance executive and attentional control (Makkar et al., 2022), DBT primarily addresses emotion regulation and impulsivity and thus, for ADHS patients, hyperactivity (Durpoix et al., 2026). In the present study, participants received a rather psychoeducational app-based intervention, which might not explicitly target emotional dysregulation to the same extent as DBT with a clinician being on-site. Nevertheless, even though the combination of tDCS and a rather psychoeducational intervention might have significantly enhanced executive functions, we did not observe differential improvement in the inattention subscale as in Basiri and Hadianfard (2023). In contrast to the complementary targeting of cognitive and affective processes reported by Basiri and Hadianfard (2023), our intervention primarily engaged cognitive control mechanisms. Further, with a total sample of 30 participants, our sample size might have been too small to detect effects particularly at the level of specific symptom subscales. Regarding the evaluation of long-term effects, the use of email-based follow-up assessments, together with the dissolution of the treatment group before follow-up, likely limited data completeness and the reliability of longitudinal measurements. Future studies should implement structured, in-person follow-up assessments and maintain group integrity throughout the course of the entire study.

With respect to **compliance**, (sham-)tDCS treatment was trackable via the Sooma portal, while the usage of the dCBT app was trackable via regular feedback from the developer of the application. Importantly, no differences in compliance for either therapy were observed between the two groups. Previous studies have suggested that additional structured contact during tDCS interventions, such as regular video calls (Dragon et al., 2024), may provide external structure that could be beneficial for individuals suffering from depression. Although additional structured contact (e.g. regular video calls) might have increased the number of completed treatment days for patients suffering from ADHD, we deliberately minimized such feedback to examine the extent to which individuals with ADHD could maintain treatment engagement through self-organization. Importantly, even with a compliance rate of approximately just 70%, participants completed a number of treatment sessions that definitely falls within the range typically applied in standard tDCS protocols for ADHD patients (e.g., around 1 – 5 sessions; Westwood, Radua & Rubia, 2021). Notably, participant feedback regarding this aspect was consistently positive, with several individuals reporting that the regular treatment sessions and associated tasks provided a helpful structure for their daily routine.

Regarding **safety**, no dropouts occurred despite the comparatively intensive treatment schedule of 60 sessions. While previous tDCS studies, e.g. for the treatment of depression, have reported participant dropouts during the intervention period due to severe side effects (Kumpf et al., 2023), here, only one participant in the present study was asked to stop the treatment prematurely by the study team after 46 sessions, with all other participants completing the protocol. Overall, these findings support the tolerability of tDCS even when applied across a relatively high number of sessions. The use of a cap with pre-punched electrode positions minimized participant-related placement errors. Further, any technical issues would have been automatically detected by the patient’s smartphone, causing the session to stop and being recorded in the Sooma portal. Additionally, the use of single-use sponges for the electrodes helped reduce the risk of skin irritation compared with repeated use of the same sponges over multiple sessions (Frank et al., 2010; Vogelmann et al., 2025). Nevertheless, we solely had to rely on patient feedback to monitor potential side effects: the participant that was asked to stop the treatment prematurely continued stimulation despite gradually developing mild itching and informed us of this only after 46 sessions. These observations underline that regular monitoring of side effects is essential to ensure participant safety throughout the study, even though treatment adherence could be tracked via a treatment platform (Vogelmann et al., 2025).

**User experience** with tDCS was rated slightly worse in the tDCS group compared to the sham-tDCS group. Given the small group sizes (15 participants per group), it should be interpreted with caution and may reflect random variation rather than a true effect of stimulation. Regarding the dCBT app, participants reported variable satisfaction. Although the dCBT app has previously been associated with significant reductions in ADHD symptom severity, our study revealed differences in patient perceptions compared to D’Amelio et al. (2026). Reductions in ASRS total scores were broadly comparable between studies, with D’Amelio et al. (2026) reporting a 5-point decrease (Cohen’s d = −.85) and the present study a 6-point decrease (Cohen’s d = −.66). However, the magnitude of perceived improvement in the present sample appeared somewhat lower than that reported by D’Amelio et al. (2026). Firstly, our cohort was younger (mean age: approximately 30 years) compared to D’Amelio’s (mean age: approximately 37). Secondly, our patients were presumably more therapy-experienced. Many were taking medication and some likely had prior psychotherapeutic treatment, which might have reduced their responsiveness to the application because of prior familiarity with CBT and mindfulness principles. Finally, the inclusion of tDCS as a secondary intervention in our study may have shifted the focus away from the dCBT app as a primary therapeutic tool. Consistent with this, response rates were low (≥30% improvement in total ASRS score: 1/15 in the tDCS group and 2/15 in the sham-tDCS group). Taken together, participants might have benefited most from the structured daily engagement, as completing the (sham-)tDCS-intervention for 20 minutes per day may have supported self-regulatory processes.

### Limitations

A major limitation of the present study is the lack of structured guidance for the use of the dCBT app, as participants were allowed to engage with the intervention at their own discretion (i.e., timing and frequency), in contrast to the (sham-)tDCS sessions, which were delivered in a standardized and daily structured manner. This discrepancy in intervention structure substantially limits the interpretability of potential additive effects and precludes firm conclusions regarding the specific contribution of the app-based component. Future studies should therefore implement a standardized protocol for the dCBT intervention, including predefined scheduling and sequencing relative to neuromodulation (e.g., fixed application days or a specified order of interventions), to enable a more rigorous evaluation of add-on effects. Further, most participants were taking ADHD medication and may have had prior psychotherapy experience, which could have influenced treatment response and complicates the attribution of effects to a third intervention. Nevertheless, at the same time, the inclusion of participants receiving standard treatments may increase the ecological validity of the findings by reflecting routine clinical practice. The relatively small sample size may have limited the generalizability of the findings. Nevertheless, given the consistently negligible effects across outcomes, it appears unlikely that increasing the sample size would have revealed a meaningful advantage of active tDCS over sham. The lack of a dCBT-only control group without tDCS further restricts conclusions about the app’s independent efficacy. While participants likely benefited from the structured daily engagement, systematic evaluation of self-organization and adherence to the dCBT app was not implemented. Furthermore, assessment of additional core domains, such as depressive symptoms or quality of life, was not included, whereas broader evaluations, as in D’Amelio et al. (2026), could provide a more comprehensive understanding of treatment effects. Finally, an intermediate assessment after six weeks could have been informative. Given the typical duration of tDCS interventions and variability in app adherence, it remains unclear whether a full 12-week intervention is necessary.

## Conclusions

This sham-controlled, randomized and single-blinded study demonstrates that home-based tDCS does not lead to further improvement in ADHD symptoms when applied as add-on to online cognitive therapy application. Importantly, the majority of patients included in the study were taking medication for ADHD. This may have modestly affected baseline symptom levels and, consequently, the observed response to an additional “third” therapeutic intervention. Future studies should include separate groups of medicated and non-medicated participants to better determine the impact of medication on treatment outcomes. Despite this limitation, the present study provides additional insight into ADHD research and contributes to the growing body of literature on non-pharmacological treatment approaches.

## Data Availability

All data produced in the present study are available upon reasonable request to the authors

## Acknowledgments

We would like to thank Dr. Andreas Kammhuber (employee at Sooma Oy, Helsinki, Finland) for organizing the 30 home-tDCS rental devices for us so quickly and bringing them to Regensburg.

